# Ceasing oxytocin in the active phase of the first stage of induced labours: A prospective audit at a tertiary hospital

**DOI:** 10.64898/2026.07.17.26358359

**Authors:** S O’Dea, G Davis, J Balendran, H Phipps, K O’Brien, B de Vries

## Abstract

**Introduction:** Oxytocin is commonly used in the process of induction of labour and is associated with uterine hyperstimulation and abnormal fetal heart rate patterns that can increase the risk of adverse perinatal outcomes. Cessation of oxytocin in the active phase of induced labour has been shown in randomised trials to reduce uterine tachysystole and abnormal fetal heart rate traces, and may reduce caesarean section. We introduced a policy recommending cessation of oxytocin infusion in the active phase of the first stage of induced labour at a tertiary hospital in Sydney, Australia, and collated both clinical outcomes and maternal satisfaction following implementation.

**Methods:** This was a prospective audit of a policy change at Royal Prince Alfred Hospital, comparing 600 women induced with oxytocin in the 6 months before the policy (November 2019–May 2020) with 556 women induced in the 6 months after implementation (June–December 2020). Eligible women had a cervix ≤5cm, an oxytocin infusion, and regular uterine contractions. The primary clinical outcome was caesarean delivery. The primary patient-centred outcome, maternal satisfaction, measured using the Six Simple Questions questionnaire, was collected in a subset of participants. Secondary outcomes included mode of birth, length of labour, uterine hyperstimulation, and perinatal outcomes.

**Results:** Caesarean delivery occurred in 29% of women before and 28% after policy implementation (p=0.77). Instrumental birth increased from 25% to 27%; and instrumental birth for maternal indications increased from 6.8% to 13% (p=0.0005). Median length of labour increased by one hour (5.4 vs 6.4 hours, p=0.006). Oxytocin was ceased for at least two hours or until birth in 13% of women before the policy versus 35% after. Maternal satisfaction scores were modestly lower after implementation (median 41 vs 38, p=0.03). Perinatal outcomes, including abnormal cord gases, Apgar scores, and NICU admission, were similar between groups.

**Conclusions:** Implementing a policy of recommending cessation of oxytocin in the active phase of induced labour did not reduce caesarean delivery rates in a real-world tertiary hospital setting, despite trial-level evidence supporting the intervention. Poor uptake, negative staff perceptions, and a modest reduction in maternal satisfaction highlight barriers to translating trial efficacy into routine clinical practice. Adequately powered trials are needed to clarify optimal protocols for oxytocin cessation and its effects on maternal and perinatal outcomes.

## Introduction

Caesarean births have been increasing globally for decades.^1^ The causes are likely multifactorial and include such factors as changing maternal demographics, changes in maternity care, medico-legal issues and maternal choice.^2–4^ Simultaneously, inductions of labour have also been increasing. In Australia, in 2023, 41% of all births were by caesarean delivery, and 33% of all births had an induction of labour.^5^ Induction of labour at term is associated with a modest reduction in caesarean delivery, and improved perinatal outcomes.^6–8^ However, oxytocin, commonly used for induction of labour, is associated with uterine hyperstimulation and fetal heart rate abnormalities that can increase the risk of hypoxic ischaemic encephalopathy and serious long-term disability.^9–11^ Interventions that reduce uterine hyperstimulation could improve these outcomes.

In one systematic review of randomised controlled trials, ceasing oxytocin in the active phase of induced labours was found to decrease the rate of caesarean delivery and decrease the rate of uterine tachysystole and non-reassuring fetal heart rate traces.^12^ However, two recent higher-quality trials included in the review found no difference for the outcome of caesarean delivery.^13,14^ Based on earlier, similar, evidence^15^ we introduced a policy of recommending cessation of oxytocin infusion in the active phase of the first stage of induced labours in a tertiary hospital in Sydney, Australia.

We audited our change in policy for relevant clinical outcomes and maternal satisfaction.

## Methods

This was a prospectively planned audit of outcomes following introduction of a policy of recommending cessation of oxytocin infusion in the active phase of the first stage of labour. This audit underwent institutional board review and received approval from the Sydney Local Health District (RPAH Zone) ethics committee (Protocol No X18-0400) on 11^th^ February 2019. It is reported using Strengthening the Reporting of Observational Studies in Epidemiology guidelines.^16^

### Setting

Royal Prince Alfred Hospital - a single tertiary hospital in Sydney, Australia with approximately 5,000 births per year.

### Population

Eligibility criteria were induction of labour using oxytocin infusion for any indication. Exclusion criteria were non-cephalic presentation, < 26 weeks gestational age and augmentation with oxytocin after labour had commenced. Labour was defined as regular painful at least 5-minutely uterine contractions with documented change in cervical dilatation.

### Intervention

The intervention was a change in policy to recommend offering ceasing oxytocin in the active phase of induced labours, outlined in Figure 1. Criteria for recommending ceasing oxytocin were (1) the labour was induced; (2) the cervix was at least 5cm dilated; and (3) there were at least 3 contractions every 10 minutes. The cervix was reassessed in two to four hours based clinical discretion and the woman’s preference. The policy was approved by the Sydney Local Health District Maternity Policy Committee and staff were educated by email notification of the new policy and recurring formal in-services. The policy came into effect on 26^th^ May 2020. The period of intervention was 13^th^ June 2020 to 13^th^ December 2020, following a wash-out period of 18 days from 26^th^ May 2020 to 12^th^ June 2020.

**Figure 1:**
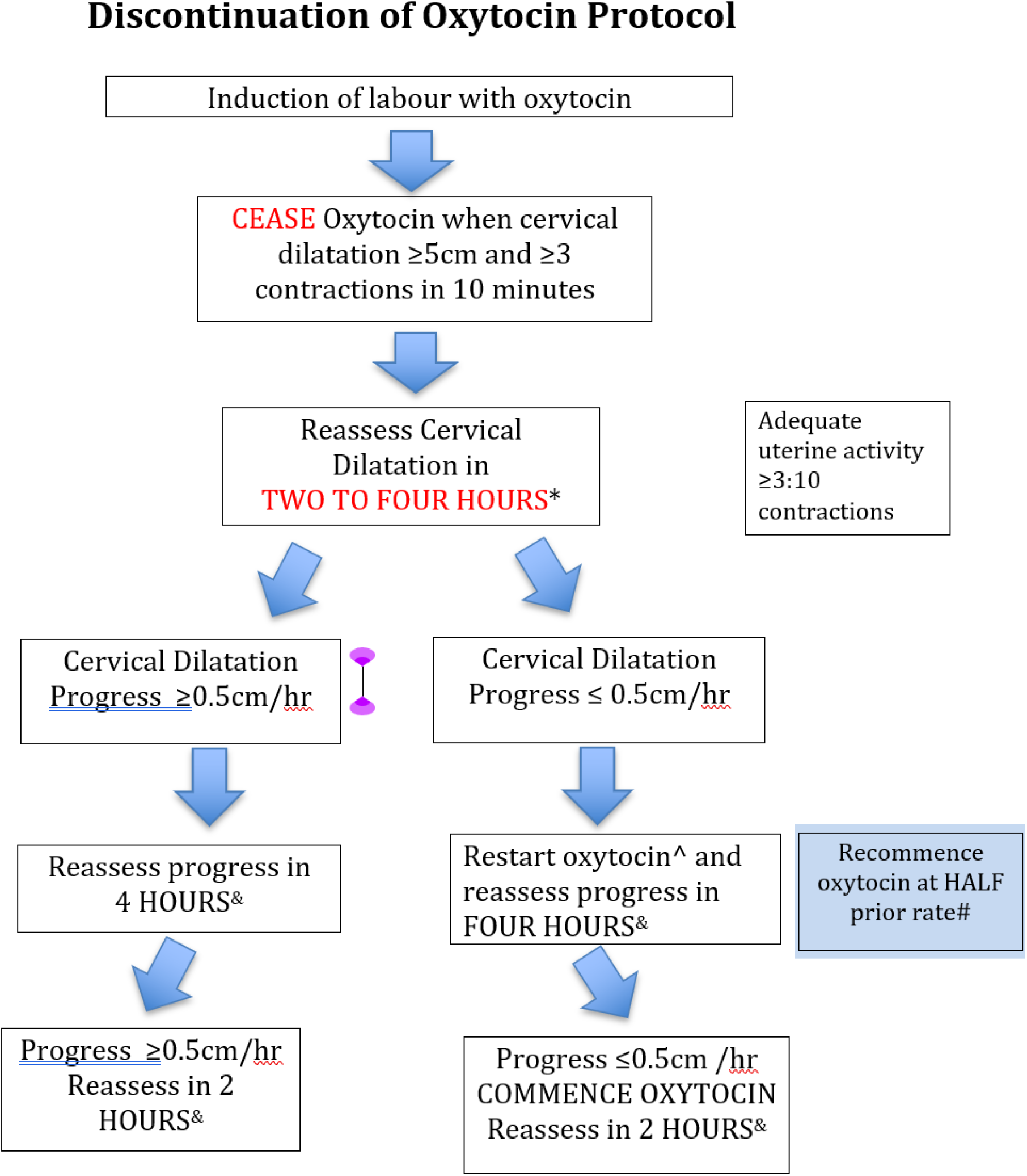
Discontinuation of oxytocin in labour protocol in a tertiary hospital in Sydney, Australia. Timing depends on clinical assessment.If ≥3:10 contractions next cervical assessmet would normally be in four hours. #HALF prior rate is rounded up (eg if ceased at 90 ml/h,recommence at 60ml/h). OXYTOCIN SHOULD NOT BE RECOMMENCED WITHOUT FIRST ASSESSING CERVICAL DILATATION &The cervix may be examined earlier if that is usual clinical practice

### Comparator

The comparison group for this audit included inductions of labour from 25th November 2019 to 25th May 2020, prior to the new policy’s implementation. The previous policy stated: “It is reasonable to consider reduction or cessation of the infusion in circumstances where spontaneous uterine activity is apparent particularly in multiparous women”. However, ceasing oxytocin in active labour was not routinely practiced.

### Data

Clinical data were collected from the electronic medical record and are routinely entered by midwives in our unit. Maternal satisfaction was recorded using a Likert scale for the question “I was satisfied with the care I received during the induction of my labour” and the six simple questions (SSQ) questionnaire.^17^ (Appendix 1) The satisfaction outcomes were collected anonymously. Maternal age was calculated to the day by subtracting the date of the mother’s birth from the date of the baby’s birth. Booking body mass index (BMI) was the measured weight on booking (usually before 20 weeks gestational age) divided by the measured height in meters squared. Public health care included all women booking through the New South Wales Health public health care system, and private health care included those who have booked through their private obstetrician or midwife. Place of maternal birth is recorded at booking.

Nulliparity was no previous births at 20 weeks or more gestational age. The main indication for induction of labour was recorded by the caring midwife. Cervical ripening was conducted using slow-release dinoprostone for 12 hours (Cervidil) (when there was no history of caesarean delivery or other uterine surgery) for a Bishop Score <7, followed by a cervical balloon (Cooks) catheter if the cervix remained unfavourable. Cervical ripening methods could be altered at the clinician’s discretion (e.g., a balloon catheter might be preferred in the presence of oligohydramnios). Gestational diabetes was diagnosed using Australasian Diabetes in Pregnancy Society criteria.^18^ Gestational hypertension was diagnosed using the 2014 Society of Obstetric Medicine of Australia and New Zealand Guideline for the Management of Hypertensive Disorders of Pregnancy.^19^ Caesarean delivery was classified according to the primary stated indication as maternal indications (generally labour arrest disorders), fetal indications (generally abnormal fetal heart rate tracings) or ‘other’. Suspected fetal compromise as an indication for induction of labour included oligohydramnios, fetal biometry (e.g., abdominal circumference or estimated fetal weight <5^th^ centile), and abnormal doppler studies (e.g., umbilical artery doppler pulsatility index (PI) >95^th^ centile, middle cerebral artery PI < 5^th^ centile, or cerebral placental ratio <5^th^ centile), or abnormal fetal heart rate tracing. Induction of labour for post-maturity occurred at 41^+4^ to 41^+6^ weeks gestational age.

### Outcomes

The primary clinical outcome was caesarean delivery. The primary patient-centred outcome was anonymously reported maternal satisfaction. (Appendix 1) Due to resource constraints, maternal satisfaction was assessed from 10^th^ March to 17th May 2020 in the pre-implementation period and 1^st^ July to 31^st^ August 2020 in the post-implementation period. Secondary maternal outcomes were unassisted vaginal birth, instrumental birth, operative birth (caesarean, forceps or vacuum) for fetal indications, the decision to restart oxytocin, ‘Red zone’ cardiotocograph according to New South Wales Health Guidelines (Appendix 2), uterine tachysystole (>5 contractions in 10 minutes, normal fetal heart rate pattern), hypertonus (contractions longer than 2 minutes or <60 seconds apart, normal fetal heart rate pattern), or hyperstimulation (tachysystole or hypertonus with abnormal fetal heart rate pattern), obstetric anal sphincter injury, use of episiotomy, epidural analgesia in labour, and postpartum haemorrhage > 1L. Secondary perinatal outcomes were abnormal arterial umbilical cord gases (pH < 7.0 or base excess < - 12mEqL^−1^), 5-minute Apgar Score <7, requirement for neonatal resuscitation, and admission to the neonatal intensive care unit. Due to concerns raised by staff during the post-implementation phase, we made a post-hoc decision to report some staff views based on two informal meetings with approximately 10 midwifery staff.

### Sample size

A sample size calculation was not performed for this clinical audit.

### Statistical methods

Normally distributed data were presented as means and standard deviations. Otherwise, medians and interquartile ranges were used. Proportions were expressed as percentages and compared using chi-squared tests. Distribution in non-parametric data were compared using the Wilcoxon rank sum test.

## Results

Of 4169 births during the audit, 1564 (37.5%) were inductions of labour and of these, 1156 (74%) used oxytocin, including 600 in the pre-implementation period and 556 in the post-implementation period. Table 1 shows baseline characteristics which were similar in the two groups. The most common indications for induction of labour were spontaneous ruptured membranes, decreased fetal movements, diabetes in pregnancy, and suspected fetal compromise.

**TABLE 1:** Baseline characteristics for 1156 participants with oxytocin induction of labour, before and after a policy of ceasing oxytocin in active labour.

|  | **Before\***<br>n = 600 | **After\***<br>n = 556 |
| --- | --- | --- |
| Age (years) | 33 (30, 36) | 32 (29, 36) |
| Pre-pregnancy BMI (kgm<sup>-2</sup>) | 24 (21, 27)<br>(2 missing) | 23 (21, 26) |
| Height (cm) | 163 (158, 167) | 164 (159, 169) |
| Place of birth |  |  |
| Australia/NZ | 260 (43) | 272 (49) |
| South East Asia | 169 (28) | 122 (22) |
| Europe | 71 (12) | 60 (11) |
| South Asia | 54 (9.0) | 59 (11) |
| Other | 46 (7.7) | 43 (7.7) |
| Public health care | 536 (89) | 503 (90) |
| Gestational age at delivery<br>(weeks<sup>+days</sup>) | 39<sup>+2</sup> (38<sup>+3</sup>, 40<sup>+3</sup>) | 39<sup>+3</sup> (38<sup>+3</sup>, 40<sup>+1</sup>) |
| Multiple gestation | 3 (0.5) | 3 (0.5) |
| IOL (as a proportion of all births) | 823/2209 (37) | 742/1960 (38) |
| Nulliparous | 450 (75) | 392 (71) |
| Oxytocin used (proportion of all IOLs) | 600/820 (73) | 556/744 (75) |
| Indication for IOL |  |  |
| SROM | 124 (21) | 117 (21) |
| DFM | 85 (14) | 72 (13) |
| Diabetes in pregnancy | 91 (15) | 81 (15) |
| Suspected fetal compromise | 89 (15) | 62 (11) |
| Post-maturity | 55 (9.2) | 37 (6.7) |
| Suspected large fetus | 36 (6.0) | 44 (7.9) |
| Hypertension in pregnancy | 30 (5.0) | 26 (4.7) |
| Advanced maternal age | 15 (2.5) | 15 (2.7) |
| South Asian Ethnicity | 3 (0.5) | 27 (4.9) |
| Choice | 9 (1.5) | 12 (2.2) |
| Other | 63 (11) | 63 (12) |
| Cervical ripening required | **333 (55)** | **334 (60)** |
| Prostaglandins only | 125 (21) | 120 (22) |
| Cervical balloon only | 82 (14) | 72 (13) |
| Both | 126 (21) | 142 (26) |
| Infant birthweight (kg)** | 3.3 (3.0, 3.7) | 3.4 (3.1, 3.7) |
| Infant sex**: Female | 311 (52) | 272 (49) |
| Male | 292 (48) | 287 (51) |

\*Data presented as median (interquartile range), mean (standard deviation), or n (%)
\*\*Three sets of twins in each group (n = 603 infants before and 559 infants after)
IOL = induction of labour

Table 2 shows maternal clinical outcomes. Caesarean delivery occurred in 29% (174/600) participants in the pre-implementation period and 28% (157/556) in the post-implementation period (X^2^ test on 1 df = 0.08; p=0.77). There were no differences in instrumental birth overall although there were more instrumental births for fetal indications in the post-implementation group (6.8% [41/600] v 13% [72/556]; X^2^ test on 1 df = 12.2; p=0.0005). The median length of labour was 5.4 hours in the pre-implementation period and 6.4 hours in post-implementation period (Wilcoxon rank sum test; p = 0.006). Figure 2 shows histograms for the lengths of labour.

**TABLE 2:** Maternal clinical outcomes for 1156 participants with oxytocin induction of labour, before and after a policy of ceasing oxytocin in active labour.

|  | Before*<br>n = 600 | After*<br>n = 556 | p |
| --- | --- | --- | --- |
| Caesarean section (total) | 174 (29) | 157 (28) | 0.77 |
| Maternal indications | 108 (18) | 91 (16) |  |
| Fetal indications | 62 (10) | 60 (11) |  |
| Other | 4 (0.7) | 6 (1.1) |  |
| Instrumental birth (total) | 149 (25) | 152 (27) | 0.33 |
| Indication: |  |  |  |
| Maternal indications | 41 (6.8) | 72 (13) | 0.0005 |
| Fetal indications | 108 (18) | 80 (14) | 0.10 |
| Type of instrument: |  |  |  |
| Forceps | 55 (9.1) | 68 (12) | 0.09 |
| Vacuum | 94 (16) | 84 (15) | 0.79 |
| Total operative birth for fetal indications | 170 (28) | 140 (25) | 0.23 |
| Oxytocin ceased |  |  |  |
| ≥ 2 hours or until birth | 80/594 (13) | 192/548 (35) | <0.0001 |
| ≥ 1 hours or until birth | 90/594 (17) | 229/548 (41) | <0.0001 |
| Oxytocin restarted** | 22/80 (27) | 108/192 (56) | <0.0001 |
| Excessive uterine activity^ | 67 (11) | 57 (10) | 0.62 |
| Abnormal CTG (at least one episode)^ | 289 (48) | 263 (47) | 0.27 |
| Length of labour (hours) | 5.4 (3.3, 8.5)<br>n = 515 | 6.4 (3.3, 9.9)<br>n = 462 | 0.006 |
| Epidural in labour | 352 (59) | 339 (61) | 0.42 |
| Perineal status |  |  |  |
| Intact | 233 (39) | 210 (38) |  |
| First degree trauma | 44 (7.3) | 36 (6.5) |  |
| Second degree trauma | 301 (50) | 291 (52) |  |
| Third/fourth degree trauma | 22 (3.7) | 19 (3.4) | 0.82& |
| Episiotomy | 160 (27) | 174 (31) | 0.08 |
| Blood loss (mL) | 350 (250, 500)<br>n = 585 | 400 (300, 600)<br>n = 528 | 0.002 |
| Postpartum haemorrhage > 1L | 28/585 (4.8) | 32/528 (6.1) | 0.35 |
| Length of stay (days) | 4.3 (3.3, 5.4) | 4.3 (3.2, 5.3) | 0.12 |

\* Data presented as median (interquartile range), mean (standard deviation), or n (%)
\*\* Where the policy's recommendation was followed (oxytocin ceased ≥ 2 hours or until birth)
^ Tachysystole, hypertonus or hyperstimulation
^^ 'Red Zone' CTG according to New South Wales Health Guidelines
& X<sup>2</sup> test for third/fourth degree tear versus no third/fourth degree tear

**Figure 2:**
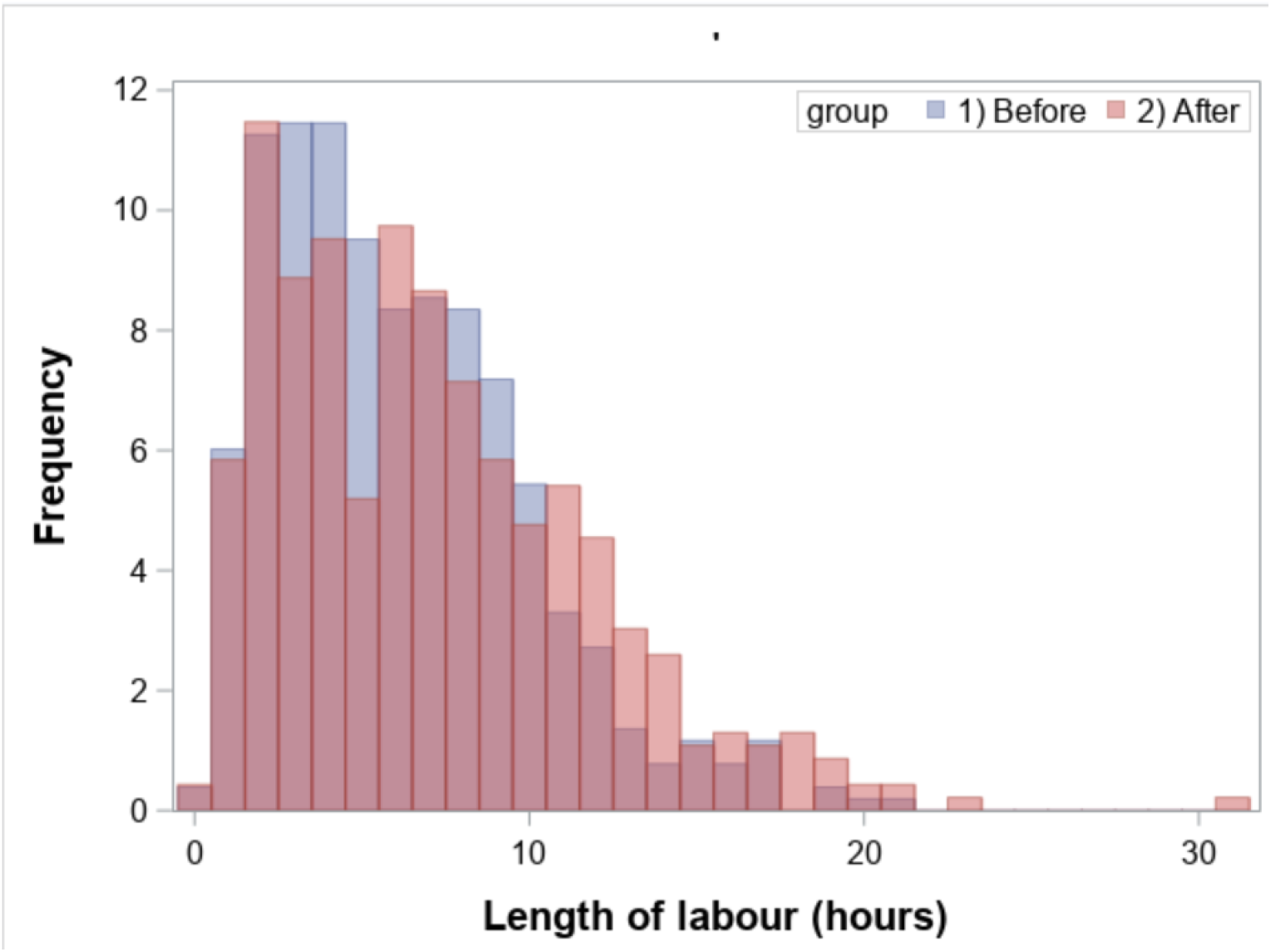
Length of labour for 977* participants with oxytocin induction of labour, before and after a policy of ceasing oxytocin in active labour. *Missing data on labour length for 179 labours

Oxytocin was ceased for ≥2 hours or until birth in 13% (80/594) participants in the pre-implementation group and 35% (190/548) in the post-implementation group.(Table 2) Of these, oxytocin was restarted in 27% and 56% respectively.

Anonymous surveys were completed by 133 participants in the pre- and 62 in the post-implementation period. The median satisfaction scores were 41 and 38 respectively (Wilcoxon rank sum test; p=0.03).(Table 2, Figure 3) Perinatal outcomes were similar in the two groups.(Table 3) Abnormal arterial cord gases were recorded in 7.0% (36/600) births and 4.9% (23/556) births in the pre- and post-implementation groups respectively. (X^2^ test on 1 df = 1.91; p=0.17)

**TABLE 3:** Maternal satisfaction for 1156 participants with oxytocin induction of labour, before and after a policy of ceasing oxytocin in active labour.

|  | Before*<br>n = 133 | After*<br>n = 62 | p |
| --- | --- | --- | --- |
| Response rate* | 133/357 (37) | 62/340 (18) |  |
| I was satisfied with the care I received during the induction of my labour | 7 (6, 7) | 6 (6, 7) | 0.006 |
| Total score ('Six Simple Questions') | 41 (37, 42) | 38 (36, 42) | 0.03 |
| (1) I had appropriate and adequate control over my care | 7 (6, 7) | 6 (6, 7) |  |
| (2) The person(s) responsible for my care were caring and compassionate | 7 (7, 7) | 6 (6, 7) |  |
| (3) Concerns that arose during pregnancy were dealt with effectively | 7 (6, 7) | 6 (6, 7) |  |
| (4) My needs were addressed with appropriate consideration for my time and individual requests | 7 (6, 7) | 7 (6, 7) |  |
| (5) The overall organisation of my care has been appropriate | 7 (6, 7) | 6 (6, 7) |  |
| (6) I would choose the same type of care for my next pregnancy | 7 (6, 7) | 6.5 (6, 7) |  |

\* n(%) or median (interquartile range) Surveys from 10<sup>th</sup> March to 17<sup>th</sup> May 2020 (pre-implementation) and from 1<sup>st</sup> July to 31<sup>st</sup> August 2020 (post-implementation).

**TABLE 4:**
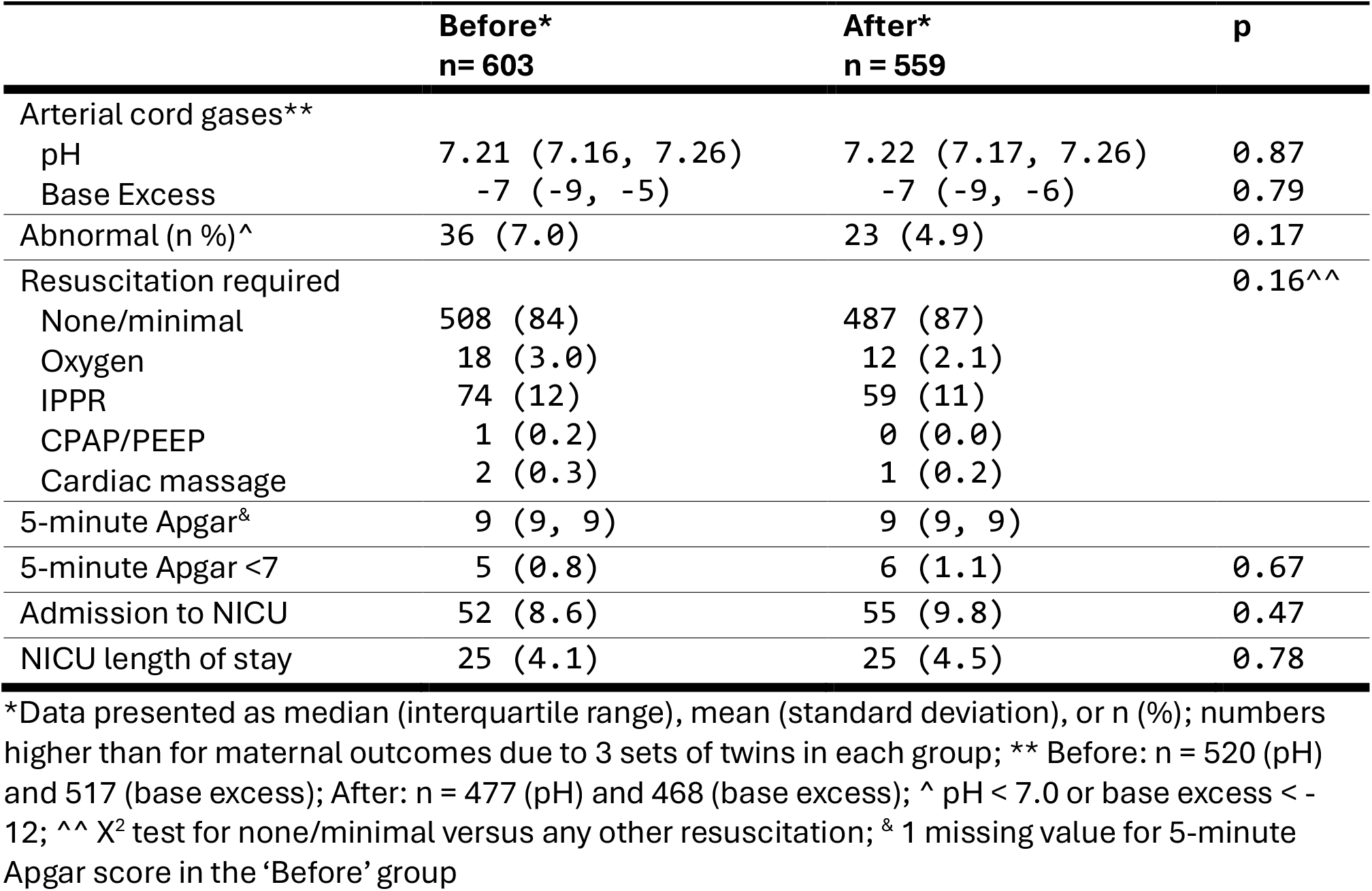
Perinatal outcomes for 1156 participants with oxytocin induction of labour, before and after a policy of ceasing oxytocin in active labour.

|  | Before*<br>n = 603 | After*<br>n = 559 | p |
| --- | --- | --- | --- |
| Arterial cord gases** |  |  |  |
| pH | 7.21 (7.16, 7.26) | 7.22 (7.17, 7.26) | 0.87 |
| Base Excess | -7 (-9, -5) | -7 (-9, -6) | 0.79 |
| Abnormal (n %)^ | 36 (7.0) | 23 (4.9) | 0.17 |
| Resuscitation required |  |  | 0.16^^ |
| None/minimal | 508 (84) | 487 (87) |  |
| Oxygen | 18 (3.0) | 12 (2.1) |  |
| IPPR | 74 (12) | 59 (11) |  |
| CPAP/PEEP | 1 (0.2) | 0 (0.0) |  |
| Cardiac massage | 2 (0.3) | 1 (0.2) |  |
| 5-minute Apgar<sup>&</sup> | 9 (9, 9) | 9 (9, 9) |  |
| 5-minute Apgar <7 | 5 (0.8) | 6 (1.1) | 0.67 |
| Admission to NICU | 52 (8.6) | 55 (9.8) | 0.47 |
| NICU length of stay | 25 (4.1) | 25 (4.5) | 0.78 |

\*Data presented as median (interquartile range), mean (standard deviation), or n (%); numbers higher than for maternal outcomes due to 3 sets of twins in each group; \*\* Before: n = 520 (pH) and 517 (base excess); After: n = 477 (pH) and 468 (base excess); ^ pH < 7.0 or base excess < -12; ^^ X<sup>2</sup> test for none/minimal versus any other resuscitation; & 1 missing value for 5-minute Apgar score in the 'Before' group

**Figure 3:**
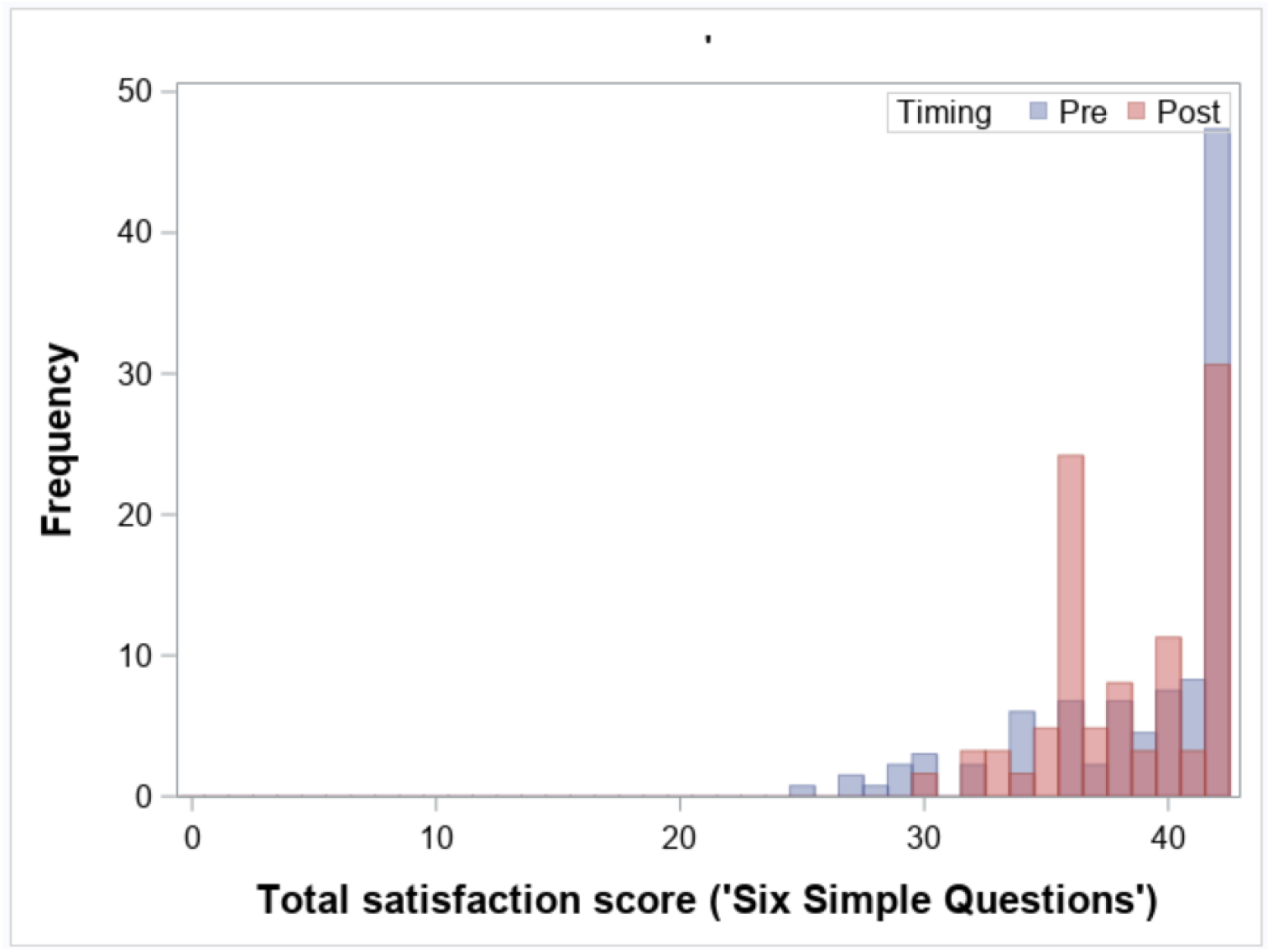
Maternal satisfaction for 195 participants with oxytocin induction of labour, before and after a policy of ceasing oxytocin in active labour.

COVID-19 restrictions were introduced progressively in New South Wales from 15 March 2020, with limits on public gatherings, followed by closure of non-essential venues from 23 March and stay-at-home restrictions from 30 March 2020. Restrictions began to ease during May and June 2020, although physical-distancing requirements and hospital visitor restrictions remained in place. Thus, the study’s pre-intervention period overlapped both the pre-pandemic period and the initial period of stringent COVID-19 restrictions, while the post-intervention period commenced during the easing phase. Hospital restrictions included limits on support people during labour and on the number and timing of visitors after birth.

Several themes emerged from post-hoc discussions with midwifery staff. These included concerns that ceasing oxytocin could cause a reduction in contractions, leading to longer labours, a risk that obstetric staff would intervene with a caesarean or operative vaginal delivery, and a perception that women wanted to continue the oxytocin. The new policy was negatively viewed by most. All staff were aware of the new policy at the time of implementation.

## Discussion

Our main observation was that a policy of recommending cessation of oxytocin in the active phase of the first stage of induced labours did not result in a decrease in caesarean deliveries. There was relatively poor uptake of the intervention, with 13% of participants having the oxytocin ceased prior to implementation of the policy compared with 35% after implementation; and a high rate of restarting the oxytocin (56%) post-implementation. There was a small decrease in maternal satisfaction from a SSQ score from 41 (mean 6.8/7) to 38 (mean 6.3/7) (Wilcoxon rank sum test; p = 0.03). This change in satisfaction scores occurred on a background of changing national restrictions during the COVID pandemic and changing local restrictions such as limits on visitation during hospital stay.

These results are important because few data are available about implementing cessation of oxytocin in active labour. While randomised controlled trials establish the efficacy of interventions under controlled conditions, barriers to implementation are common, where clinician behaviour, adherence and organisational factors may influence outcomes. Midwifery staff did not view ceasing oxytocin favourably, due to concerns about slowing labours and increasing the risk of caesarean delivery, although caesarean delivery rates were not impacted by the policy (29% before compared with 28% after implementation). The rate of restarting oxytocin post-implementation of 56% was higher than reported in randomised controlled trials which reported overall rates of about one-third with wide variation from four to 41%^12^. This may reflect staff beliefs and behaviour. In one randomised trial in Denmark, the authors reported:

“*Anecdotally, birth attendants involved with the trial reported a general impatience with slower than average rates of cervical dilatation, leading to pressure from medical and midwifery colleagues to restart oxytocin even though the trial criteria for this had not been met*”.^13^

We observed an increase in operative vaginal delivery for maternal indications from 6.8% to 13% (X^2^ = 12.2; p = 0.001) an outcome that was not reported in the systematic review by Whitley et al (2025), although they did observe that overall operative vaginal delivery for any indication was similar when oxytocin was ceased compared to when it was continued.^12^ An increase in operative vaginal delivery for maternal indications is physiologically plausible because less oxytocin could reduce the strength or frequency of contractions leading to a longer second stage and consequent intervention with forceps or vacuum birth. However, this explanation is speculative for this secondary outcome, as there is risk of a Type 1 error due to multiple tests of statistical significance, and a high risk of bias inherent in observational studies with historical controls.

We additionally observed an increase in the median length of labour by one hour, which is consistent with the 30-minute increase reported the systematic review by Whitley et al.^12^ The small increase in median blood loss of 50mL is of uncertain clinical significance and may be a Type I error given no difference was observed in postpartum haemorrhage in the randomised trials.^12^

Induction of labour at term is known to result in a modest decrease in caesarean delivery,^6,8^ and there is evidence that this effect could be mediated through preventing caesarean delivery for slow progress rather than for fetal distress or intrapartum asphyxia/hypoxia.^20^ Consequently, reducing fetal distress in induced labours could result in an even greater reduction in caesarean deliveries. Uterine hyperstimulation is a well-recognised complication of oxytocin which can in turn lead to umbilical cord compression and fetal hypoxaemia. Thus, any intervention that decreases the dose or need for oxytocin has the potential to improve perinatal outcomes including intrapartum stillbirth and neonatal hypoxic ischaemic encephalopathy (HIE).^21,22^ Globally, intrapartum stillbirth causes 900,000 deaths per year,^23^ and there are more than one million cases of HIE.^24^

A strength of this study is that the intervention was evaluated in a real-world clinical setting, allowing assessment of its effectiveness under routine conditions in which clinician and patient behaviours, adherence, and organisational factors may influence outcomes. Another strength was assessment of maternal satisfaction using a validated questionnaire.

Weaknesses included the before-and-after study design, because changes unrelated to the intervention could account for any observed changes over time. For example, changes in maternal satisfaction could be related to the COVID-19 epidemic. Another weakness was the low response rate for maternal satisfaction, likely related to increased staffing requirements during the pandemic, with clinical staff having limited time for clinical research, and the anonymity of the survey.

Future research should focus further on cessation of oxytocin in active labour because there is strong evidence there is a reduction in uterine tachysytole, associated with fetal distress due to umbilical cord compression; and strong evidence of a reduction in abnormal fetal heart rate traces in labour, in turn associated with HIE and long-term neurological dysfunction.^25^ There is a need for adequately powered randomised controlled trials, particularly in low-income settings where intrapartum hypoxia is more common, and to determine the safest way to induce labours. For example, the ideal target number of contractions, frequency of titration and continuous versus pulsatile administration of oxytocin remains unknown. These options would be particularly suited to a large multinational adaptive platform trial where multiple management options can be assessed more efficiently than in a traditional randomised trial.

## Conclusions

Implementing a policy of recommending cessation of oxytocin in the active phase of induced labour did not reduce caesarean delivery rates in a real-world tertiary hospital setting. Poor uptake, negative staff perceptions, and a modest reduction in maternal satisfaction indicate barriers to translating trial efficacy into routine clinical practice.

## Data Availability

All data produced in the present work are contained in the manuscript

## Appendix 1

### RPA Women and Babies Induction of Labour Satisfaction Survey

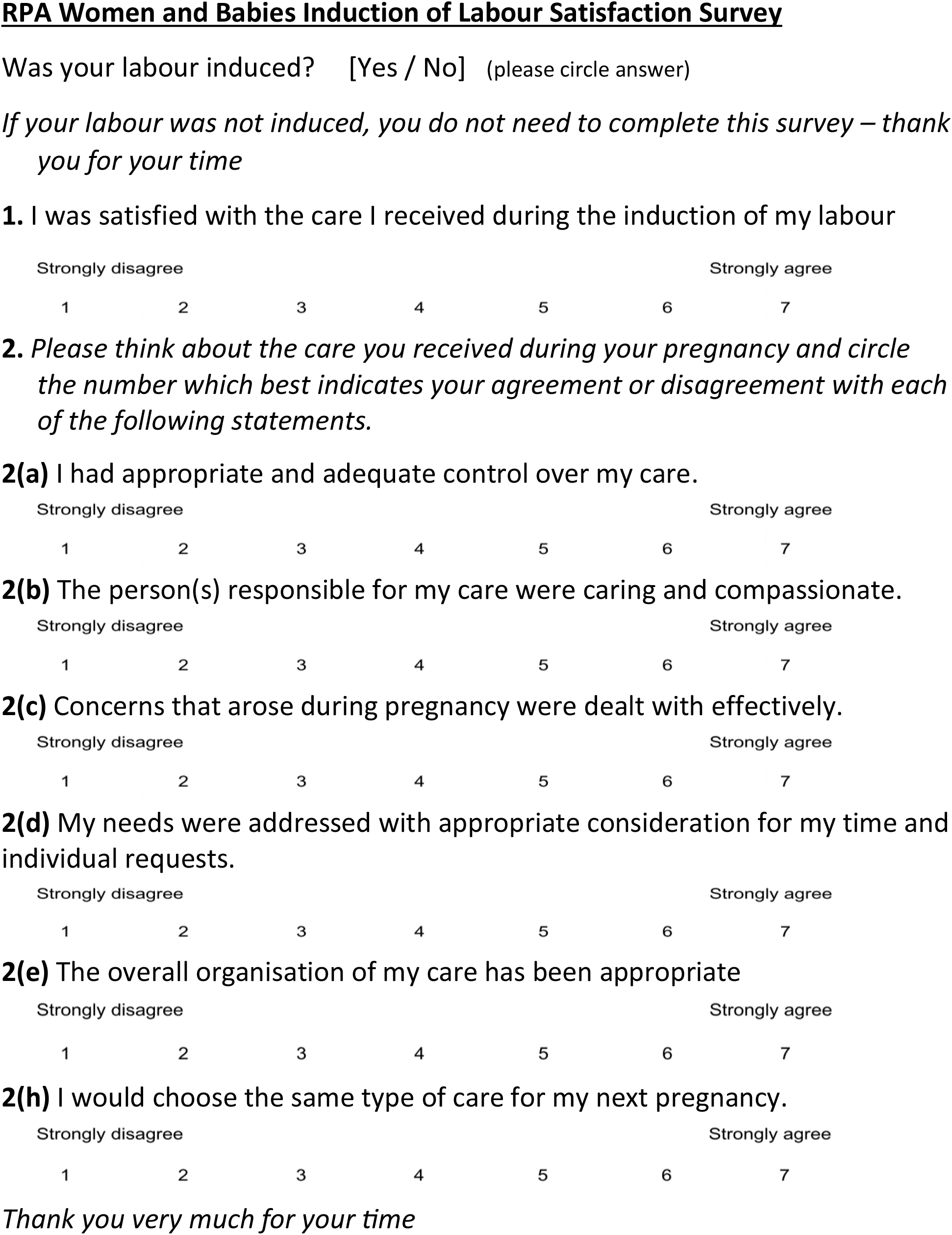

| 1 | 2 | 3 | 4 | 5 | 6 | 7 |
|---|---|---|---|---|---|---|

| 1 | 2 | 3 | 4 | 5 | 6 | 7 |
|---|---|---|---|---|---|---|
| 1 | 2 | 3 | 4 | 5 | 6 | 7 |
| 1 | 2 | 3 | 4 | 5 | 6 | 7 |

## Appendix 2

### New South Wales Health Intrapartum Monitoring Algorithm

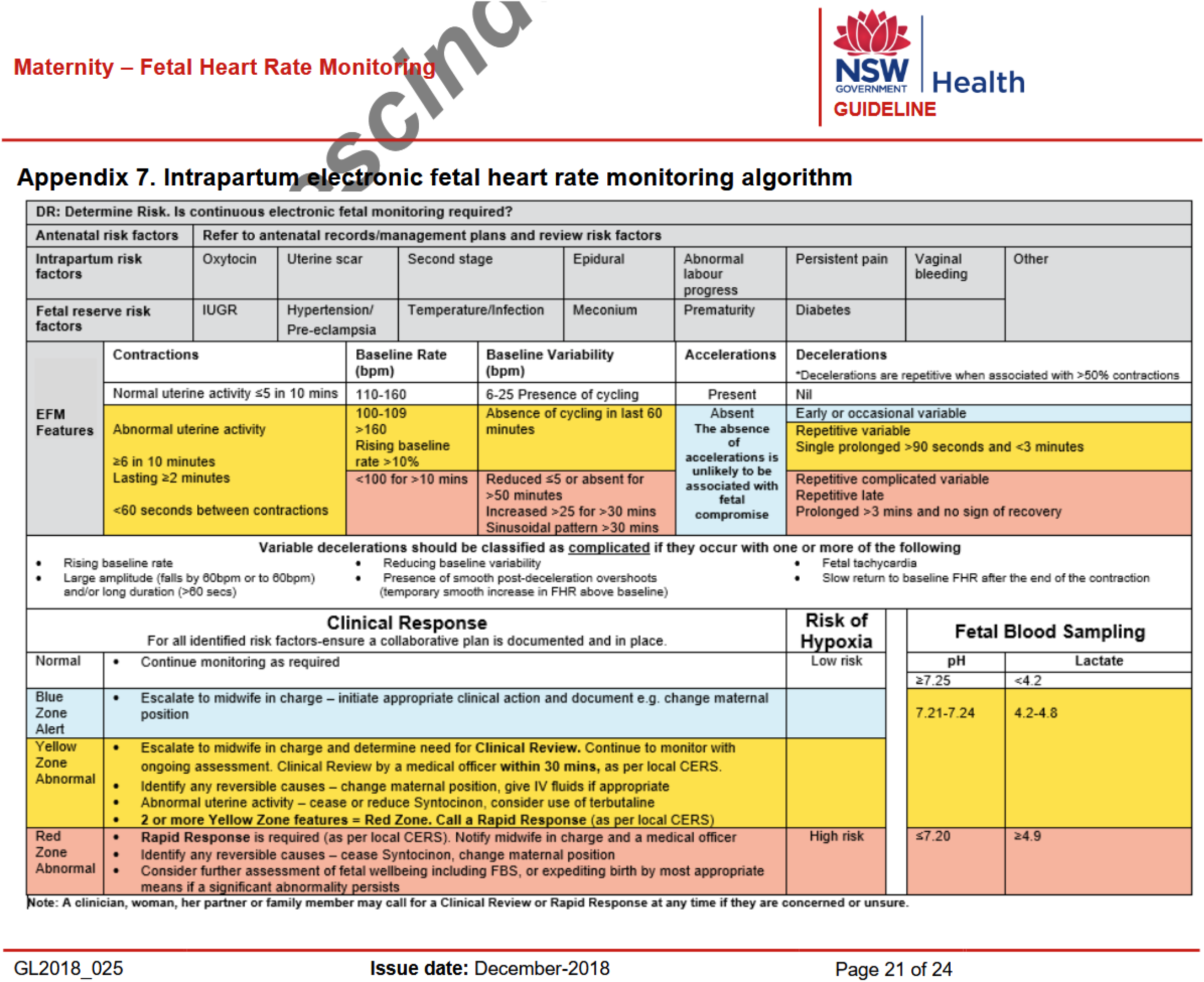

